# Are the upper bounds for new SARS-CoV-2 infections in Germany useful?

**DOI:** 10.1101/2020.07.16.20155036

**Authors:** Wolfgang Bock, Thomas Götz, Yashika Jayathunga, Robert Rockenfeller

## Abstract

At the end of 2019, an outbreak of a new coronavirus, called SARS–CoV–2, was reported in China and later in other parts of the world. First infections were reported in Germany by the end of January and on March 16th the federal government announced a partial lockdown in order to mitigate the spread. Since the dynamics of new infections started to slow down, German states started to relax the confinement measures as to the beginning of May. As a fall back option, a limit of 50 new infections per 100,000 inhabitants within seven days was introduced for each city or district in Germany. If a district exceeds this limit, measures to control the spread of the virus should be taken. Based on a multi– patch SEAIRD–type model, we will simulate the effect of choosing a specific upper limit for new infections. We investigate, whether the politically motivated bound is low enough to detect new outbreaks at an early stage. Subsequently, we introduce an optimal control problem to tackle the multi–criteria problem of finding a bound for new infections that is low enough to avoid new outbreaks, which might lead to an overload of the health care system, but is large enough to curb the expected economic losses.

## 1. Introduction

Since its first appearance in Wuhan, China, in December 2019, SARS-CoV-2 became a threat worldwide and imposed massive challenges to different societies see e.g. [1]. The virus is currently, as of July 9th 2020, attested in every country, with a total of 12,068,034 detected cases and 550,159 associated deaths [2]. There are various models predicting massive outbreaks, if the countries do not slow down a further spread by taking countermeasures [3, 4, 5].

In the absence of vaccines and reliable pharmaceutical treatment, non– pharmaceutical interventions (NPIs) were established in almost every country and are since the objective of intensive studies [4, 6]. First infections were reported in Germany by the end of January [7, 8] and on March 16th the federal government announced a partial lockdown in order to mitigate the spread [9]. Despite hints for the efficacy of NPIs, there is growing criticism from the economical point of view due to unemployment, challenges for companies, and a possible threat for the social system. A fall of income by around 70% together with a loss of consumption is discussed [10]. For Germany, a loss in GDP of 10% per week for severe NPIs is forecasted [11]. In rapid succession, out–of–lockdown strategies and dynamical NPI strategies were proposed all around the world [6]. The German government, specifically, imposed upper bounds for the number of newly infected on the level of districts as a quantitative criterion to assess if a lockdown, including school and shop closure, would be locally necessary [9]. The upper bound was set to 50 new infections per 100,000 inhabitants within seven days on the level of individual German districts (Landkreise), see [12]. Districts in which this bound is passed, should enforce severe NPIs for this particular district, but also provide the possibility for the inhabitants to enter the surroundings for shopping and restaurant visits. The upper limit of 50 newly infected per 100,000 citizens seems not to be based on scientific considerations, but rather on political interests.

In this article, we use a multi-patch-SEAIRD-type model to evaluate the spread of COVID-19 in the German state of Rhineland-Palatinate with its 36 districts. We use an optimal control problem to find upper and lower bounds for which a lockdown can be invoked or eased in each individual district. At this, mobility within the districts plays a key role. To introduce mobility into the system, a residence-time-budgeting matrix was computed based on the commuting data of the districts [13]. As proposed by the German government, the bounds for all districts are assumed to be equal. For the corresponding cost functional, we penalize (i) the time in lockdown, (ii) the number of switches from and into lockdown, to mimic the negative effects on the economy, as well as (iii) the number of casualities.

## 2. Methods

### 2.1. Study design, source of data and study settings

We used a multi-patch-SEAIRD-model to describe the COVID-19 transmission dynamics under time-dependent NPIs for the districts of the German state Rhineland-Palatinate. Since the German government has invoked a rule for the NPIs valid for all districts of Germany, we assumed that this strategy will be also valid for new upper and lower bounds of the newly infected in one week. Altogether, we made the following assumption for our study. 1) The case data in the districts, obtained by the Johns-Hopkins University, were correct up to the most recent date of this study. 2) The commuting data of Rhineland-Palatinate [13], represent the mobility within the districts suitably. 3) Workplace situation already has reached pre-pandemic normality. 4) Model parameters can be adopted from a previous parameter analysis study for COVID-19 [14], which had been based on time series for Germany. 5) Data on age and comorbidities could be neglected for this study.

### 2.2. Modelling NPIs

We considered two different transmission scenarios, namely inner- and outer-household transmissions. While the transmission within households hardly preventable, NPIs can lead to a significant reduction of the outer-household contacts and hence reduce the outer-household transmissions [3]. In SIR models, this effect can be achieved by reducing the basic reproduction number, see e.g. [6, 15, 16]. For an uncontrolled spread of COVID-19, a reproduction number around 2.6 was found, while post-intervention studies showed a value of 0.62 for certain European countries [16], which corresponds to a reduction of 74.5%. In particular in the case of Germany a reproduction number of 2.2 was deduced from data in [14]. With no detailed intervention scheme available, we modelled this effect by a reduction of the transmission rate. The period in which the NPIs should be valid in each district were obtained by solving an optimal control problem, in which the absolute time of the lockdown, the deaths from COVID-19 and the number of switches to lockdown and back within a period of 200 days were penalized. The latter represents the economical challenge for a company or a district, respectively, to implement these interventions: weekly on-off strategies, with changing measures on a short-term basis are certainly not desirable.

### 2.3. Multi–Patch–SEAIRD–type model for the regional spread of COVID-19

The mathematical model, which was used throughout this paper constitutes an extension of the model from [14], including the spatial spread of the disease due to movements within the population. This model is a compartmental model of ordinary differential equations in which the total population is divided into six compartments, namely: Susceptible *S*_*i*_, Exposed *E*_*i*_, Asymptomatic *A*_*i*_, Infected *I*_*i*_, Recovered *R*_*i*_ and Deceased *D*_*i*_, where the index corresponds to the specific patch *i*. The state of Rhineland–Palatinate, with a total population of about 4 million inhabitants, is divided into 24 districts and 12 independent cities. Hence, we considered a total of 36 different patches (district or city) and end up with the 216–dimensional ODE–system:

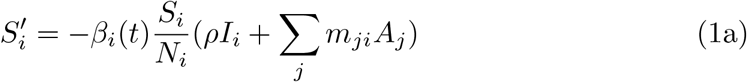

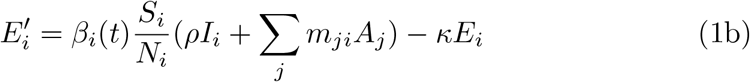

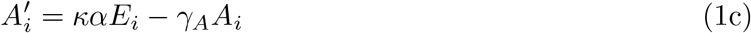

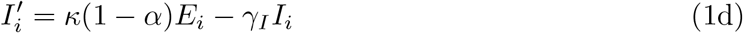

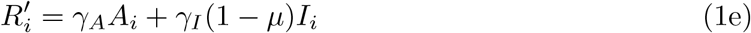

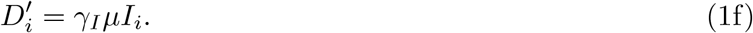

Here *β*_*i*_(*t*) denotes the time-dependent transmission rate in patch *i*; due to possible reinforcement of restrictions within individual patches, the transmission rate can vary in time and between districts. Hence, the transmission rate in district *i* was assumed to contain the following three components:

1. A multiplicative factor *u*_*i*_(*t*) ∈ (0.2, 1], modelling the control via restrictions to public and economic life. The unrestricted case corresponds to *u*_*i*_ = 1 with smaller values of *u*_*i*_ representing more severe restrictions.
2. A base value 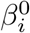, representing an average transmission rate within the patch.
3. A uniform distributed centered random variable 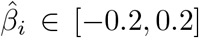, allowing to capture random fluctuations of *±*20% of the outbreaks in different districts.

Summarizing, the transmission rate writes as

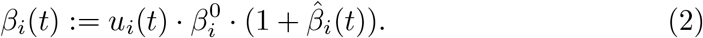

The mobility of healthy and asymptomatic infected people, who are often not even aware of being infectious, is described by a so-called residence-budgeting-time matrix *M* = (*m*_*ji*_). It is important to note that 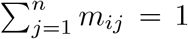, where *m*_*ij*_ ∈ [0, 1]. The term *m*_*ij*_ in residence-time budgeting matrix refers to the fraction of time spent in another patch [17]. Based on the commuting matrix for Rhineland-Palatinate, hence the average mobility behaviour within the state, the matrix *M* can be deduced, by assuming that an individual spends 1*/*3 of the day at work (at the state it commutes to if not working in its residing state) and 2*/*3 of the day in its residing state. For symptomatic infected, we assume no mobility. The incubation period equals 1*/κ* and *α* denotes the fraction of infected being asymptomatic. Recovery periods for asymptomatic and symptomatic equal 1*/γ*_*A*_ ≤ 1*/γ*_*I*_ and finally *µ* denotes the death rate for symptomatic cases. For asymptomatic cases, lethality is neglected.

The values for the parameters used in model (1) were taken from [14] and are listed in Table 1.

**Table 1:**
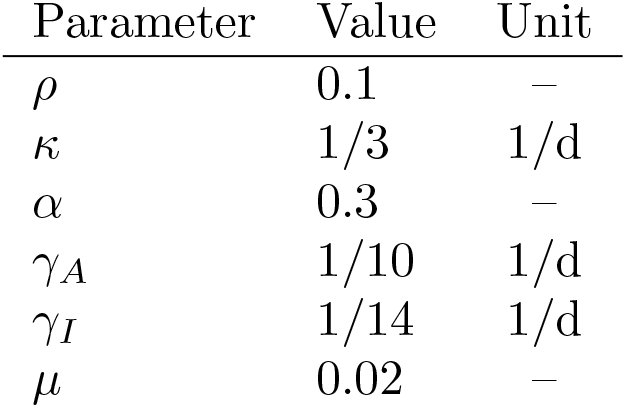
Parameter values of model (1).

The target variables (*Z*_*i*_)_*i*=1,*…,M*_ of the model represent the new infections per 100, 000 inhabitants during the last seven days within a district. This observable had been defined by the German government. Computing the number of new infections for a time span of 7 days, we obtain

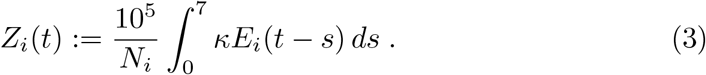

We embedded the question of meaningful upper bounds for new infections into an optimal control framework. The state-wide upper bounds *Z*_min_ and *Z*_max_ serve as the optimization variables to be determined. If the target variable *Z*_*i*_ in a district exceeds the bound *Z*_max_, a re-implementation of restrictions in the given district is invoked. These restrictions are relaxed again if the target variable drops below a lower bound *Z*_min_. The two bounds *Z*_max_ and *Z*_min_ which were assumed to be constants for every state, serve as the optimization variables to be determined.

For the multiplicative factor *u*_*i*_(*t*), controlling the transmission rate, we obtain

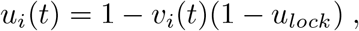

where *u*_*lock*_ *<* 1 denotes the reduction of 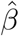 due to restrictions. The indicator *v*_*i*_(*t*) switches between 0 (no measures) and 1 (lockdown)

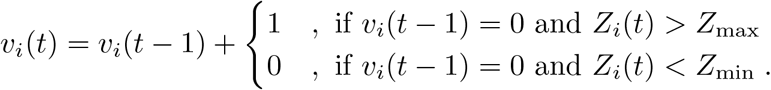

Here, we assumed that reimposed restrictions lead to an 80% reduction of the transmission. This corresponds to a slightly stricter reduction as found in [16].

The objective of the control problem takes three aspects into account

1. The economic loss due to reimposed restrictions. According to [11] the shut–down costs per month account for approx. 40% of the GDP, i.e. 10% per week.
2. The number of fatalities *D*_*i*_.
3. The number of switches *Q*_*i*_ into the shut down, since it can be expected that there is an economic loss by imposing the measures within the district, thus making a continuous on-off strategy not feasible.

Hence, we propose the following cost functional as objective of the minimization problem

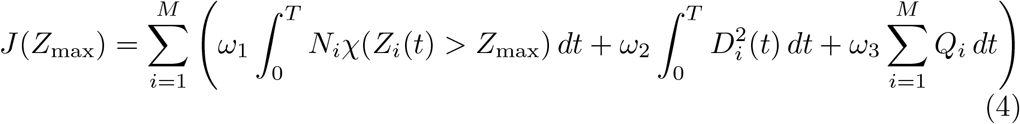

where *χ* denotes the characteristic function, i.e. *χ*(*Z*_*i*_(*t*) *> Z*_max_) = 1, if *Z*_*i*_(*t*) *> Z*_max_ and zero otherwise. The weights are set to *ω*_1_ = 1000, *ω*_2_ = 1 and *ω*_3_ = 5 in order to balance the different terms in the cost functional.

For the simulation of system (1), we used a standard explicit Euler method. The optimal control problem, i.e. the search for optimal parameters *Z*_min_ and *Z*_max_ as lower and upper bounds for the target variable *Z*, was solved with the built-in MATLAB-routine `fminsearch`. Note that the existence of a minimizer is ensured due to the convexity of the cost functional.

The time-dependent transmission rate is a random variable, which has to be taken into account when comparing the cost functional for different parameters *Z*_min_ and *Z*_max_. For this purpose, a static random seed of 1000 sample paths of 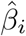, individually for every district, was used. This seed was also used when comparing scenarios with and without the influence of mobility, wherein the latter the residence-time-budgeting matrix corresponds to the identity.

Figure 1 displays the dependence of the weighted duration of restrictions and the number of deaths in Rhineland-Palatinate as a function of the upper bound *Z*_max_ of new infections within 200 days and for different lower bounds *Z*min.

**Figure 1:**
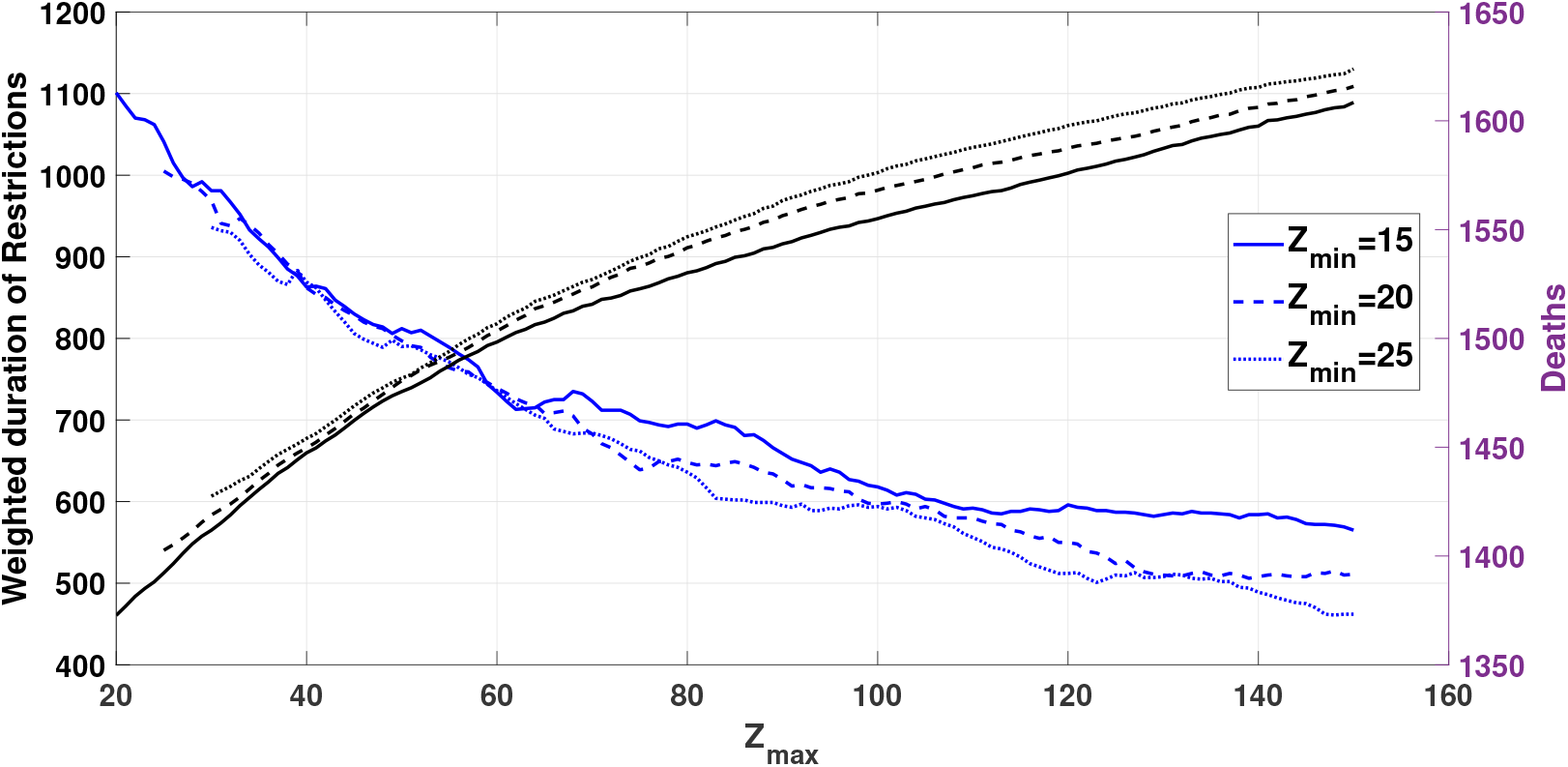
Weighted duration of restrictions in man-days (blue) and deaths (black) as a function of *Z*_max_ and *Z*_min_ for whole Rhineland-Palatinate. The different line styles indicate the lower bound *Z*_*min*_ = 15 (solid), 20 (dashed) or 25 (dotted).

As expected, the total number of man-days in lockdown, summed up over all districts, is monotonically decreasing with growing *Z*_max_ for all values of *Z*_min_. Note that small oscillations occur due to the time restriction of the simulation, which was chosen to be 200 days. Naturally, the duration of the restrictions increases with larger intervals between the upper and lower bound. The number of deaths is increasing with growing *Z*_max_ and growing *Z*_min_ respectively. One can clearly see the non-linear behaviour in both curves.

## 3. Results

### 3.1. Demographic characteristics and data of COVID–19 cases

Table A.2 in the Appendix presents the population size, case numbers, recovered inhabitants and the death numbers for each district in Rhineland-Palatinate as of July 6, 2020. We assume a dark figure of active cases which is equal to the tested active cases and add 10 % are asymptomatic and 10% are exposed cases as initial conditions. Since at the time of the study 37% of the total ICU beds in Germany was free, see [7] (12065 free ICU beds for 305 COVID-19 patients with intensive care), the capacity of the health care system in the districts is neglected.

### 3.2. Simulation and predicted optimal intervention strategy

#### 3.2.1. Disease dynamics

In case of no control being imposed, there will be outbreaks in the districts leading to 6708 deaths per 100,000 within the time horizon of 200 days. Mobility plays a role at the beginning of the spreading. Figure 3 and 2 show the disease progression in 4 different characteristic districts, once with and once without mobility between the districts. The *R*_0_-value throughout the simulation in the non-controlled phase was 2.2, which is in accordance with previous findings [14]. Note that in this scenario, the initial conditions are based on the current data, i.e. there are cases in different districts already. The mobility hence accelerates the spreading. In Neuwied in Figure2 infections occur, due to the fact that the district is disease-free. The imports of infections from other districts lead to a later outbreak in Neuwied compared to the other districts displayed. One can see one more effect of the mobility in Figure 3 in the case of Neuwied. While in the other displayed districts the curve for the uncontrolled number of infections (red) is below the *Z*_max_ value in a good agreement with graph of the controlled one until the first peak, both graphs differ slightly for Neuwied. This is due to the fact that the control of the other districts also plays a role on the disease progression of Neuwied in a scenario where mobility is concerned.

**Figure 2:**
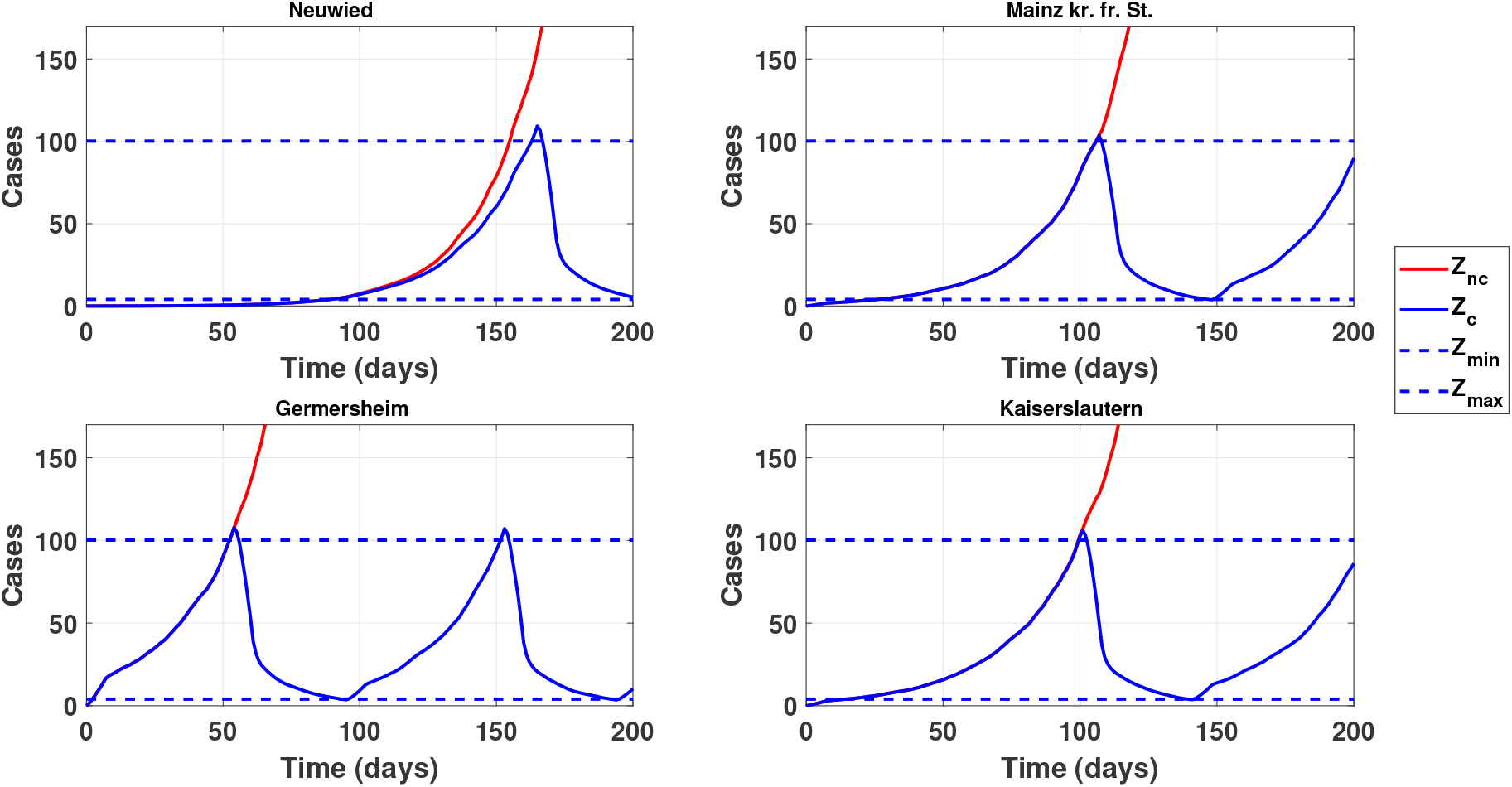
Controlled and uncontrolled disease progression for four districts of Rhineland-Palatinate including mobility between the districts, with *Z*_min_ = 4 and *Z*_max_ = 100.

**Figure 3:**
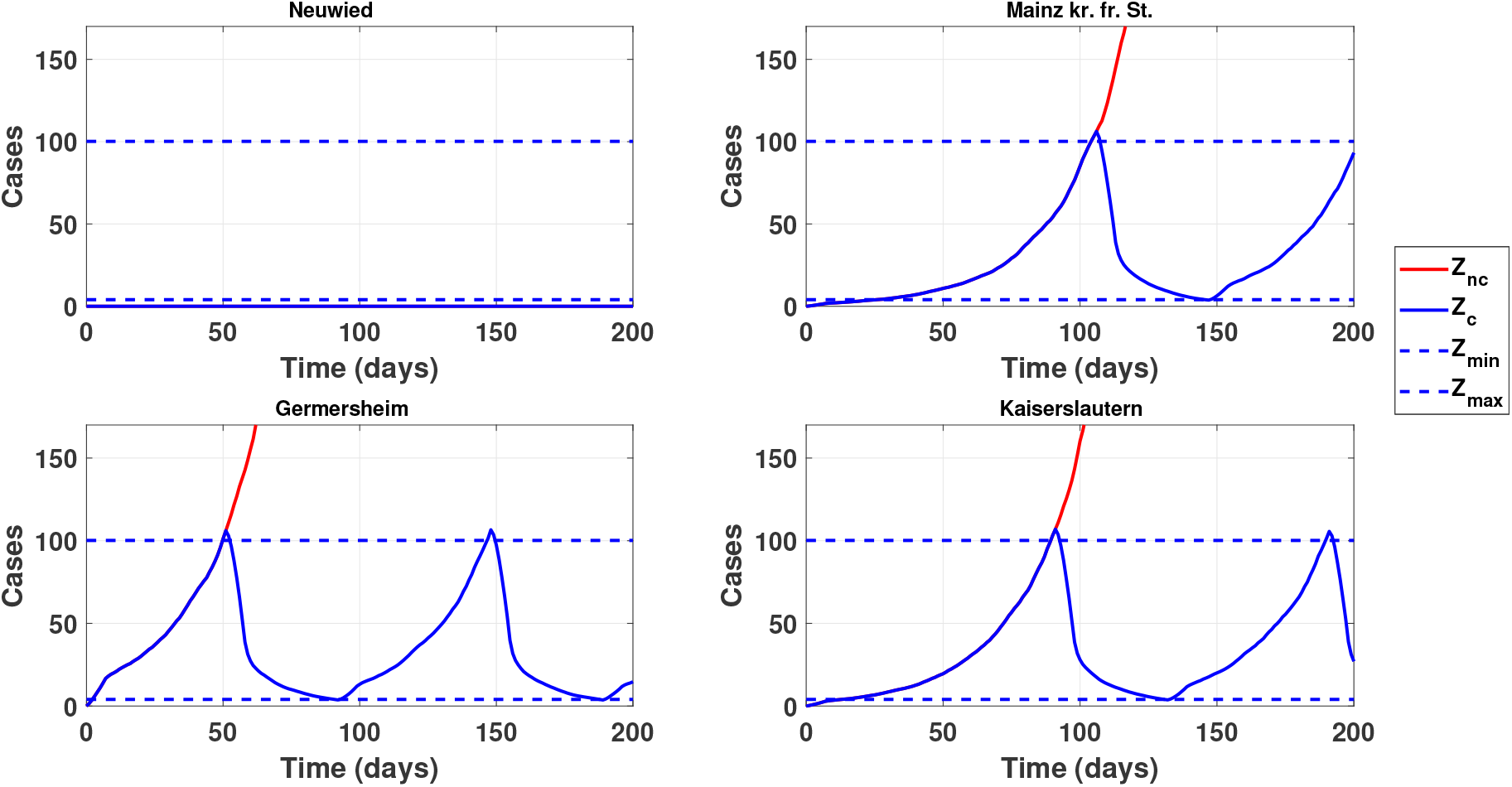
Controlled and uncontrolled disease progression for four districts of Rhineland-Palatinate without mobility between the districts, with *Z*_min_ = 4 and *Z*_max_ = 100.

#### 3.2.2. Optimal bounds for the number of newly infected per week

Based on our model, we formulated an optimal control problem where the number of death, the time of restrictions and the number of switches from a non-NPI to an NPI regime is minimized subject to the underlying multi-patch system. The time of the restrictions here was weighted with the population of the district to incorporate the economical importance of the district within the state. Based on 1000 random samples, the lower and upper bounds were optimized w.r.t. the mean of the above costs, using `fminsearch`. As optimal upper and lower bounds, we found *Z*_max_ = 100 and *Z*_min_ = 4. A sample path of *Z* with these values is displayed in Figure 5. A sample path for the infected number of cases per 100,000 inhabitants with these values is displayed in Figure 4. We see that indeed the number of infected can effectively controlled by the NPIs.

**Figure 4:**
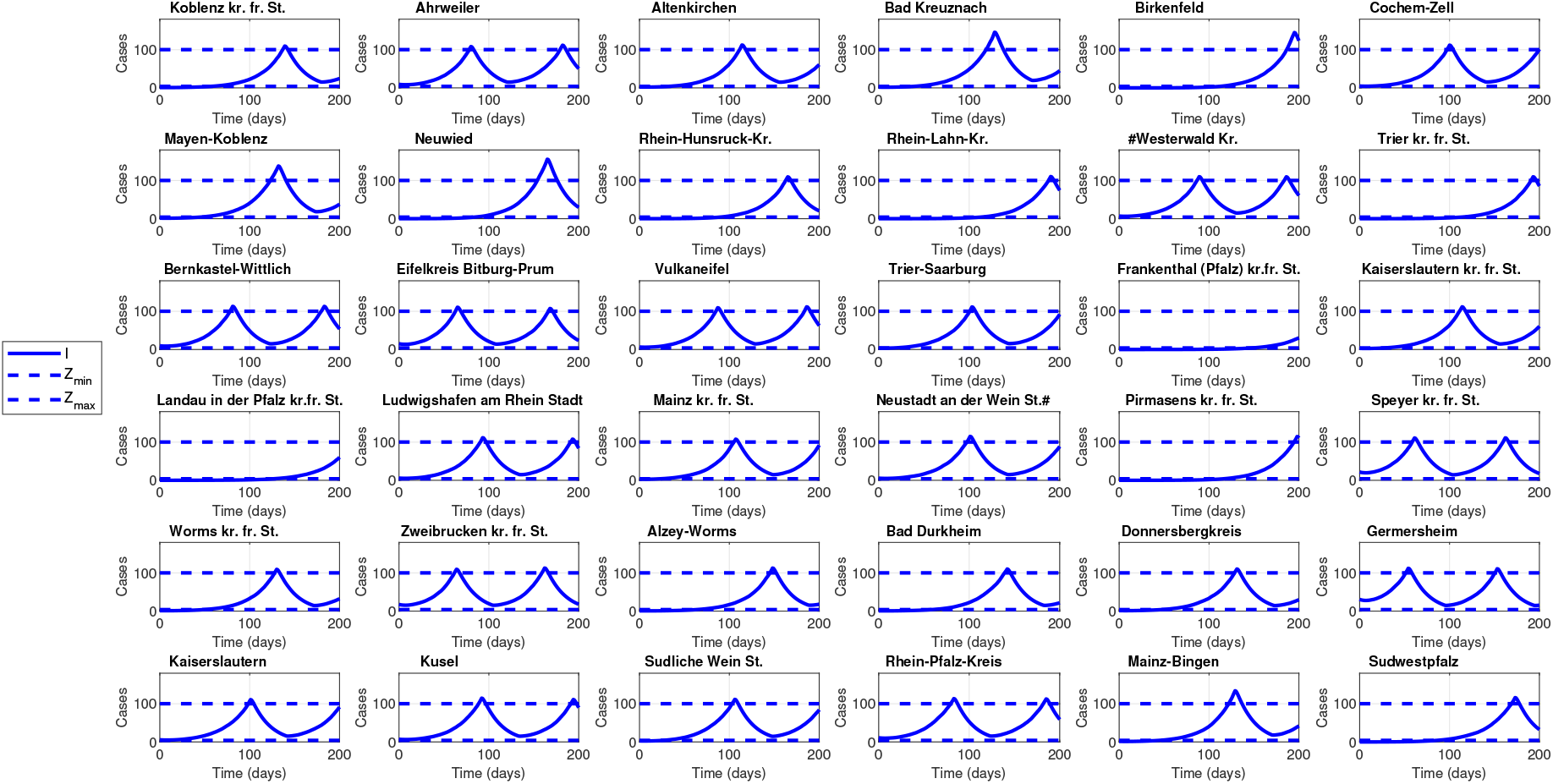
A sample path for the infected number of cases per 100,000 inhabitant with the optimized values obtained for the upper bound and lower bound, *Z*_max_ = 100 and *Z*_min_ = 4.

**Figure 5:**
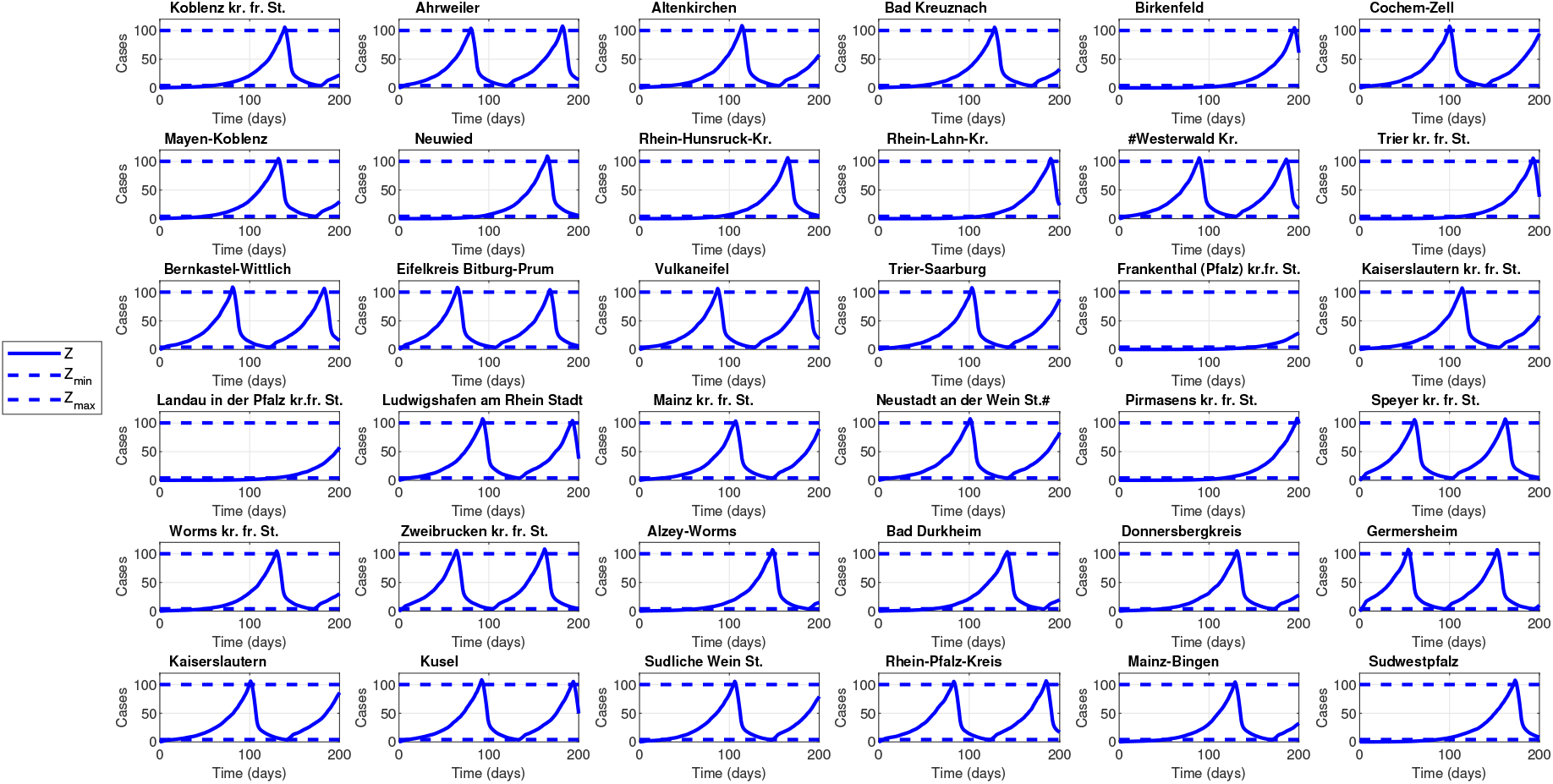
Number of newly infected over a 7 days timespan with controlled disease progression for *Z*_max_ = 100 and *Z*_min_ = 4.

Overall, mobility here had only a minor contribution to the disease progression. Without mobility, there are few districts with no active disease cases at day 0 and thus no control is needed over the full-time horizon. Including mobility, these districts can become infected by imports and control measures, i.e. NPIs, have to be taken. However in districts where there were already infected cases at the initial time, the mobility results just in a slight time shift in the time series of infected.. Figure 6 illustrates the multiplicative factor *u*_*i*_(*t*), controlling the transmission rate over the time considered with the two scenarios including and excluding mobility effect. There are several counties such as Frankenthal (Pfalz) kr.fr. St. and Landau in der Pfalz kr.fr. St. in which the control is not switching over time as the number of newly infected over a 7 days time span is not exceeding the upper limit during the considered 200 days. All the other counties are having a switch once or two times during the considered time span accordingly to the number of newly infected over a 7 days time. For disease-free districts such as e.g. Neuwied and Trier kr.fr. Stadt a lockdown is just invoked in the case of mobility and at a very late timepoint.

**Figure 6:**
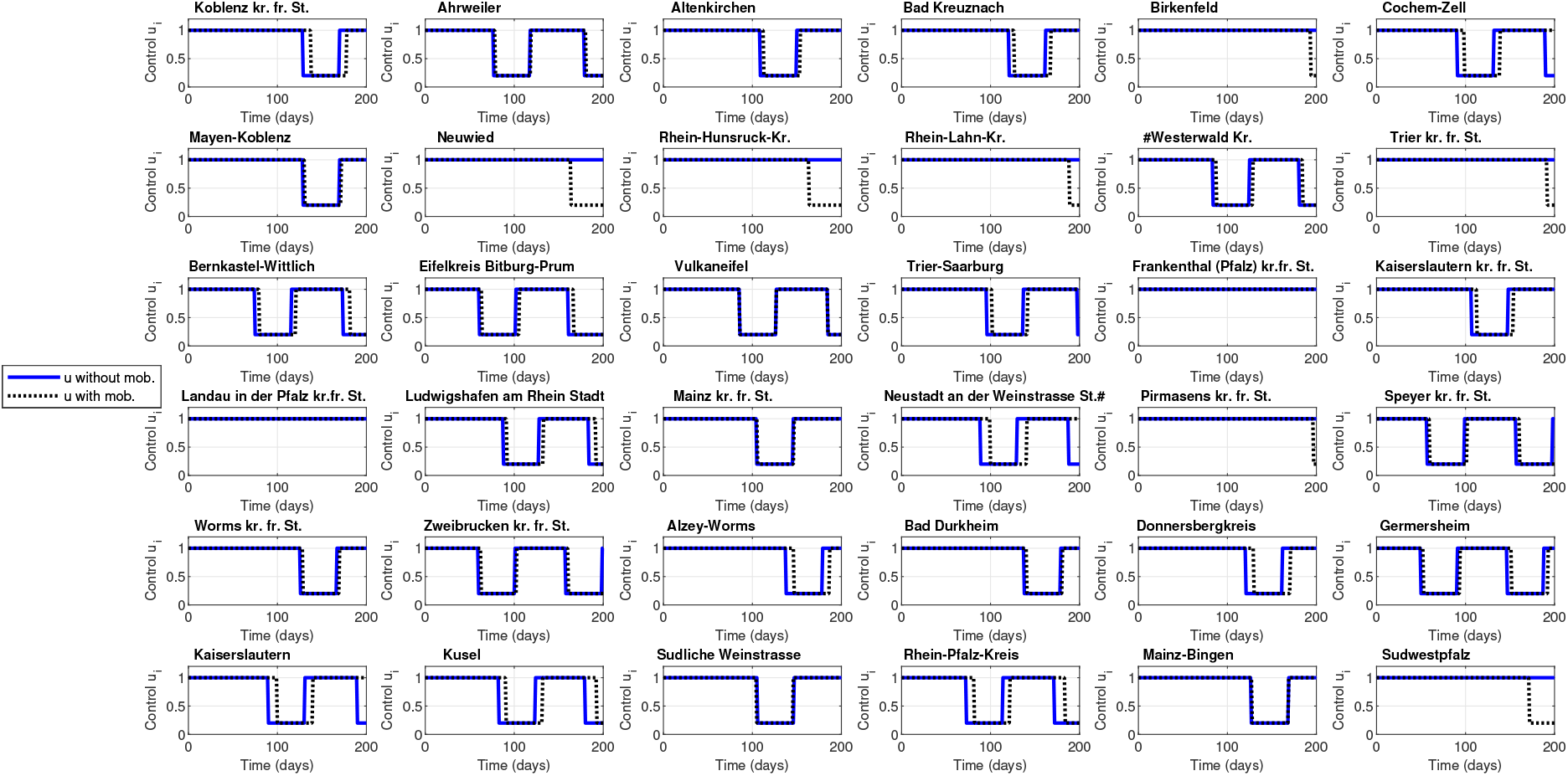
Time dependent control for for *Z*_max_ = 100 and *Z*_min_ = 4.

## 4. Discussion

In this study, we used a multi-patch-SEAIRD-type model with an optimal control problem to deduce optimal bounds for the minimal and maximal number of new infections per week, in order to ease or invoke NPIs as countermeasures for the COVID-19 spreading. The optimal values were computed based on the disease dynamics in the 36 districts of the German state Rhineland-Palatinate. To include random changes in the contacts and to obtain robustness, we perturbed the transmission rate with a uniformly distributed random variable and performed the optimization over a fixed seed of 1000 random samples of the transmission rate.

Our study has several findings. First, we showed that the mobility between the districts plays a minor role if the disease is already spread in the whole state. However, disease-free districts can become infected due to imports. Second, comparing the controlled and the non-controlled scenarios, a small number of NPI phases, which last around 40 days, lead to effective control of the disease progression. The amount of newly infected in 100,000 per week is strictly higher than the 50 suggested by the German government. The lower bound is also strictly larger than 0. Note that quantitatively the number is of course related to the weights set in the optimization problem. For the weights in our scenario the control is realizable and the disease can effectively be kept under control with at maximum 2 lockdowns in 200 days.

We assumed here the dark figure of infected cases to be as high as the number of detected cases,. i.e. the number of infected at the beginning of the simulation is really able to contribute to new infections. The number in Table A.2 reflects however the detected cases are under quarantine and are hence not infecting other susceptibles. Backtracking and effective testing, together with household quarantine, can, of course, influence our result positively. Thus, our study can be seen as a conservative estimate. We neglected many heterogeneities in the population and omitted super-spreading events and comorbidities, which can, of course, influence the results. Our study may have important implications since we displayed findings relevant for decision-makers in the fight against COVID-19. We provide a strategy that can be easily adapted and used to control the disease progression in Rhineland-Palatinate effectively. The measures can be seen as dynamic since they are adapted to the number of cases in the districts and hence work adaptive to their specific economic needs and also the needs of the whole state. Further, the findings stimulate further research such as the influence of the household structure, which seems crucial for the progression [3], the influence of reduced mobility, i.e. is it really necessary to shield a district completely if the case numbers are not yet high. The influence of basic patient data such as comorbidities, hospitalization rate and the influence of testing and backtracking will certainly lead to more realistic results.

## Data Availability

All data or supplementary material is accessible openly and all sources are referred to in the manuscript.

## Appendix A. Demographic and disease status of the districts of Rhineland-Palatinate

**Table A.2:**
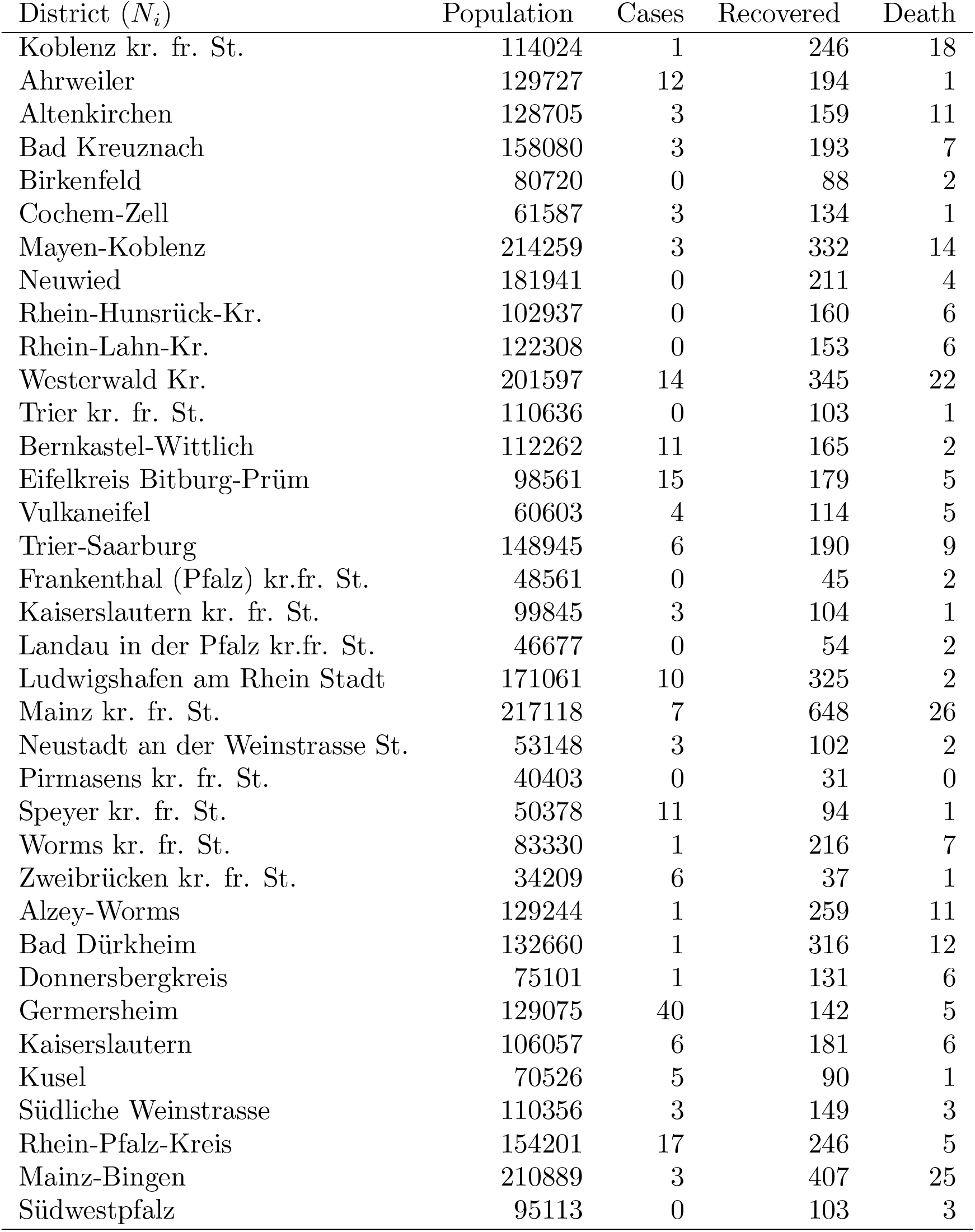
The data source for the population is the Statistisches Bundesamt Deutschland. The data source for the number of cases is Robert-Koch-Institut (RKI), Berlin. The number of cases, recovered and death data areas as of July 6, 2020.

| District ($N_i$) | Population | Cases | Recovered | Death |
| --- | --- | --- | --- | --- |
| Koblenz kr. fr. St. | 114024 | 1 | 246 | 18 |
| Ahrweiler | 129727 | 12 | 194 | 1 |
| Altenkirchen | 128705 | 3 | 159 | 11 |
| Bad Kreuznach | 158080 | 3 | 193 | 7 |
| Birkenfeld | 80720 | 0 | 88 | 2 |
| Cochem-Zell | 61587 | 3 | 134 | 1 |
| Mayen-Koblenz | 214259 | 3 | 332 | 14 |
| Neuwied | 181941 | 0 | 211 | 4 |
| Rhein-Hunsrück-Kr. | 102937 | 0 | 160 | 6 |
| Rhein-Lahn-Kr. | 122308 | 0 | 153 | 6 |
| Westerwald Kr. | 201597 | 14 | 345 | 22 |
| Trier kr. fr. St. | 110636 | 0 | 103 | 1 |
| Bernkastel-Wittlich | 112262 | 11 | 165 | 2 |
| Eifelkreis Bitburg-Prüm | 98561 | 15 | 179 | 5 |
| Vulkaneifel | 60603 | 4 | 114 | 5 |
| Trier-Saarburg | 148945 | 6 | 190 | 9 |
| Frankenthal (Pfalz) kr.fr. St. | 48561 | 0 | 45 | 2 |
| Kaiserslautern kr. fr. St. | 99845 | 3 | 104 | 1 |
| Landau in der Pfalz kr.fr. St. | 46677 | 0 | 54 | 2 |
| Ludwigshafen am Rhein Stadt | 171061 | 10 | 325 | 2 |
| Mainz kr. fr. St. | 217118 | 7 | 648 | 26 |
| Neustadt an der Weinstrasse St. | 53148 | 3 | 102 | 2 |
| Pirmasens kr. fr. St. | 40403 | 0 | 31 | 0 |
| Speyer kr. fr. St. | 50378 | 11 | 94 | 1 |
| Worms kr. fr. St. | 83330 | 1 | 216 | 7 |
| Zweibrücken kr. fr. St. | 34209 | 6 | 37 | 1 |
| Alzey-Worms | 129244 | 1 | 259 | 11 |
| Bad Dürkheim | 132660 | 1 | 316 | 12 |
| Donnersbergkreis | 75101 | 1 | 131 | 6 |
| Germersheim | 129075 | 40 | 142 | 5 |
| Kaiserslautern | 106057 | 6 | 181 | 6 |
| Kusel | 70526 | 5 | 90 | 1 |
| Südliche Weinstrasse | 110356 | 3 | 149 | 3 |
| Rhein-Pfalz-Kreis | 154201 | 17 | 246 | 5 |
| Mainz-Bingen | 210889 | 3 | 407 | 25 |
| Südwestpfalz | 95113 | 0 | 103 | 3 |

## Appendix B. Supplementary figures

**Figure B.8:**
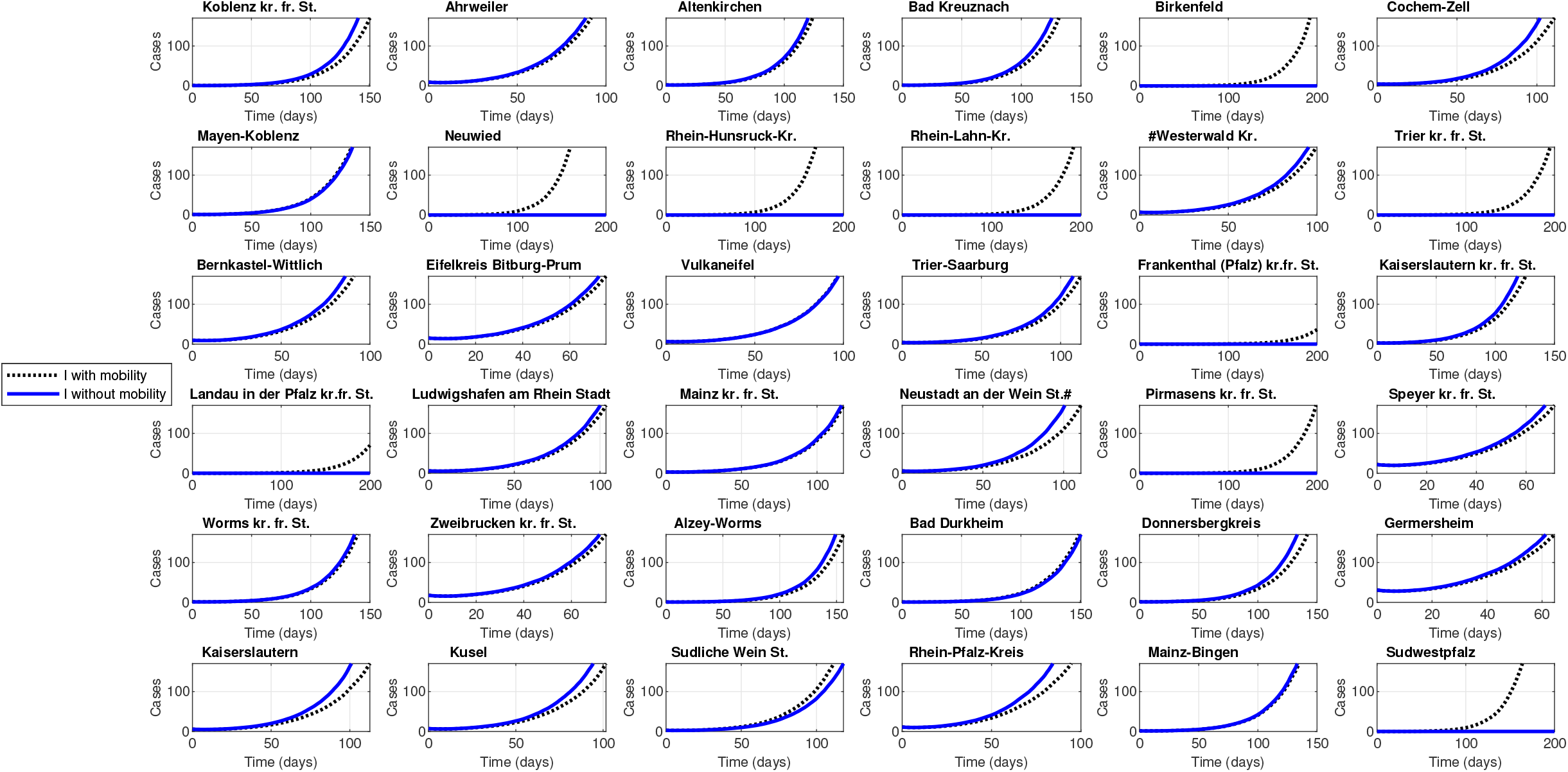
Simulation with no control (*u*_*lock*_ = 1): Infected cases with and without mobility over time in the 36 counties of RLP. The results are scaled to 100; 000 inhabitants per county.

**Figure B.9:**
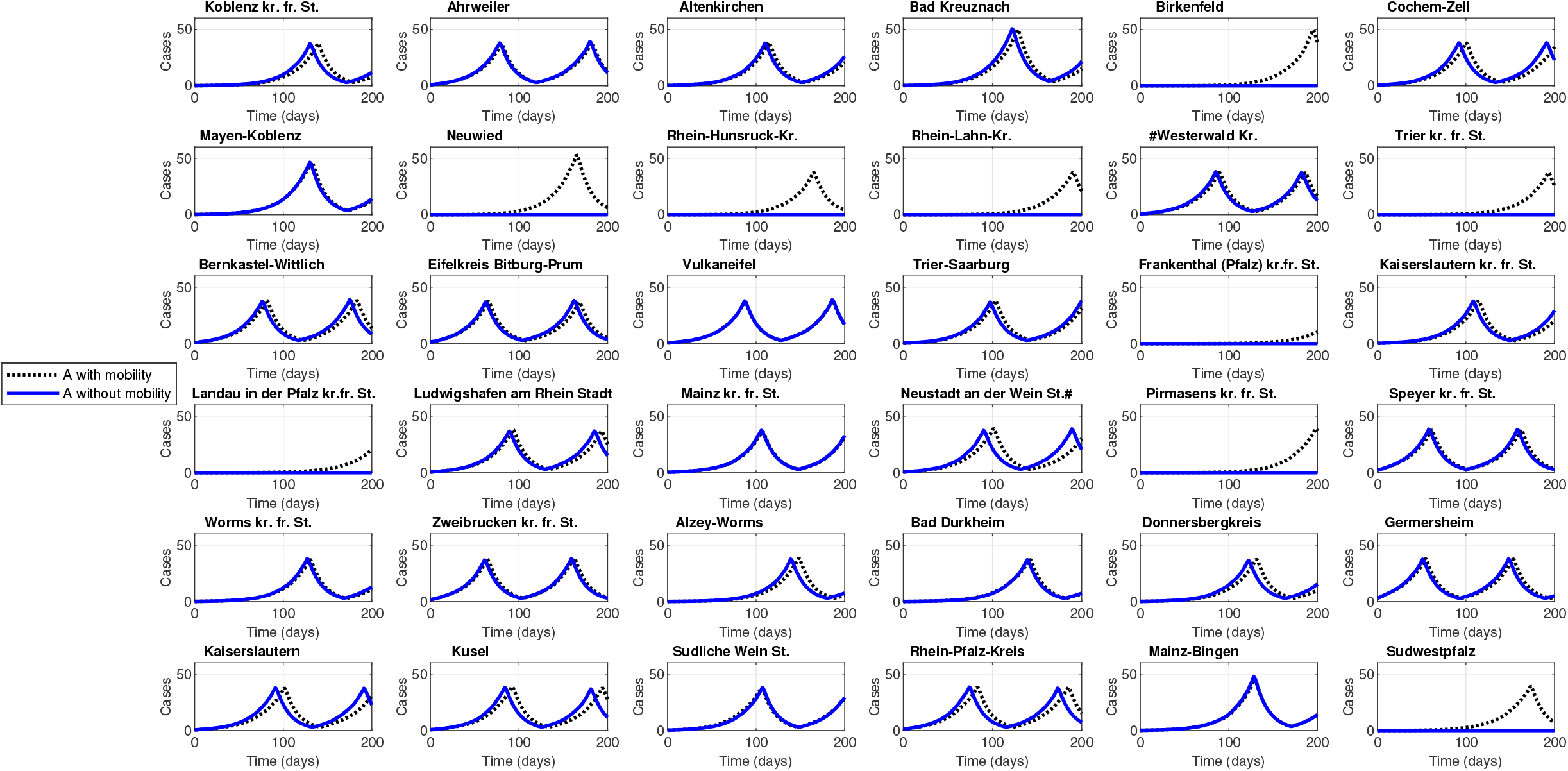
Simulation with control (*u*_*lock*_ = 0.2): Asymptomatic cases with and without mobility over time in the 36 counties of RLP. The results are scaled to 100; 000 inhabitants per county.

**Figure B.10:**
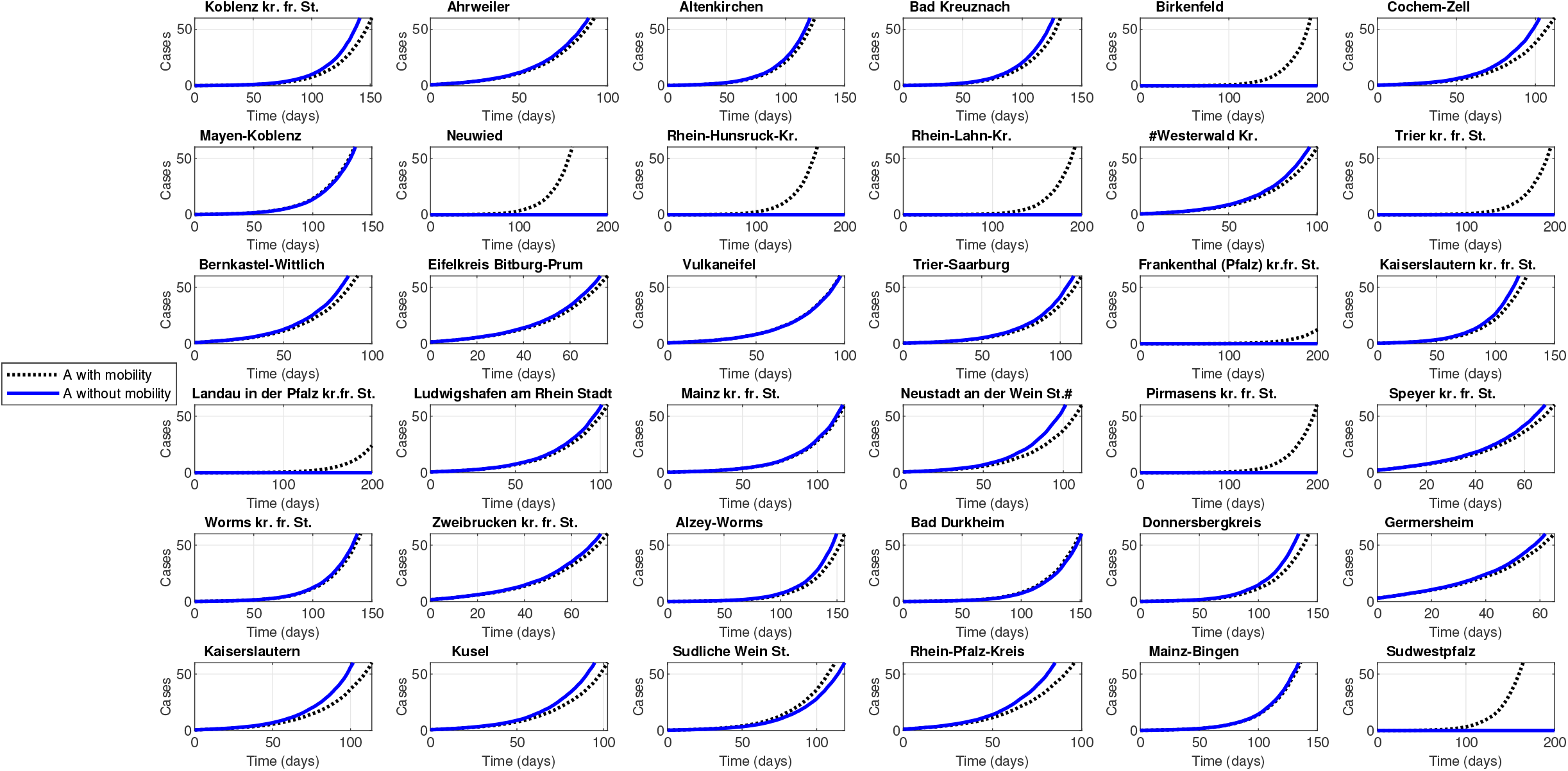
Simulation with no control (*u*_*lock*_ = 1): Asymptomatic cases with and without mobility over time in the 36 counties of RLP. The results are scaled to 100; 000 inhabitants per county.

**Figure B.11:**
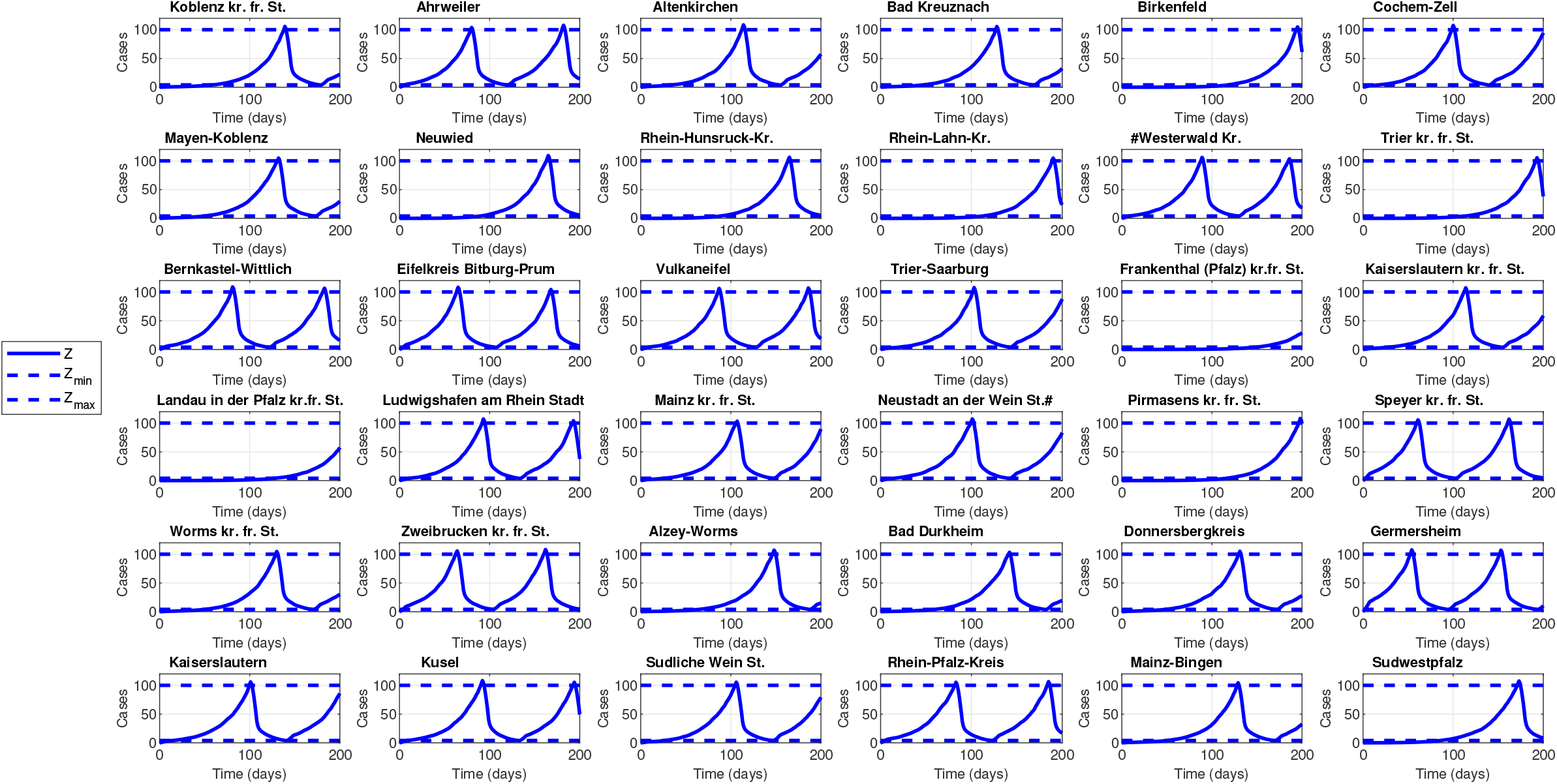
Simulation with no control (*u*_*lock*_ = 1) and with mobility: New cases in the past 7 days per 100; 000 inhabitants in the 36 counties of RLP.

**Figure B.12:**
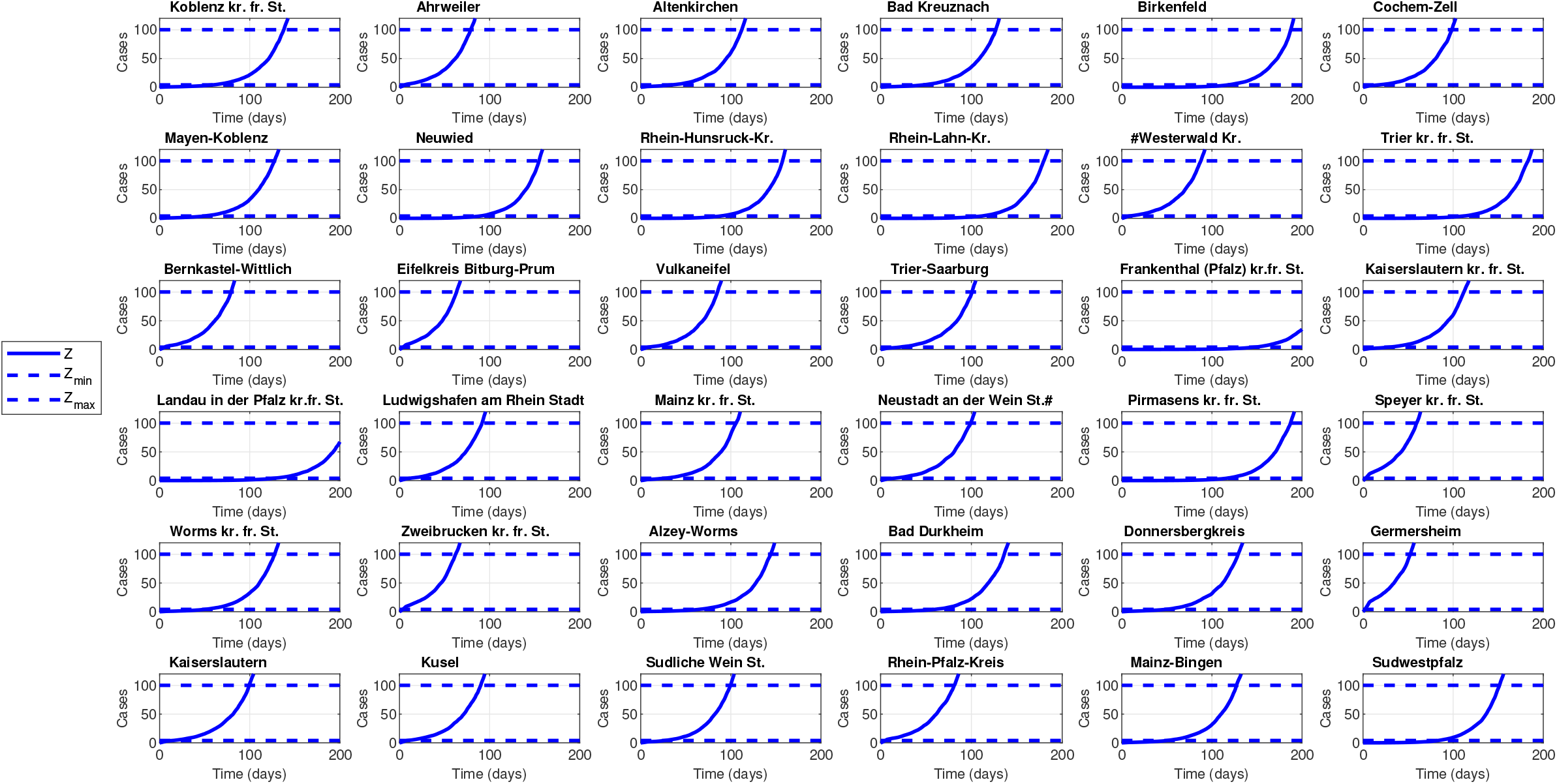
Simulation with no control (*u*_*lock*_ = 1) and with mobility: New cases in the past 7 days per 100,000 inhabitants over time in the 36 counties of RLP with mobility.

**Figure B.13:**
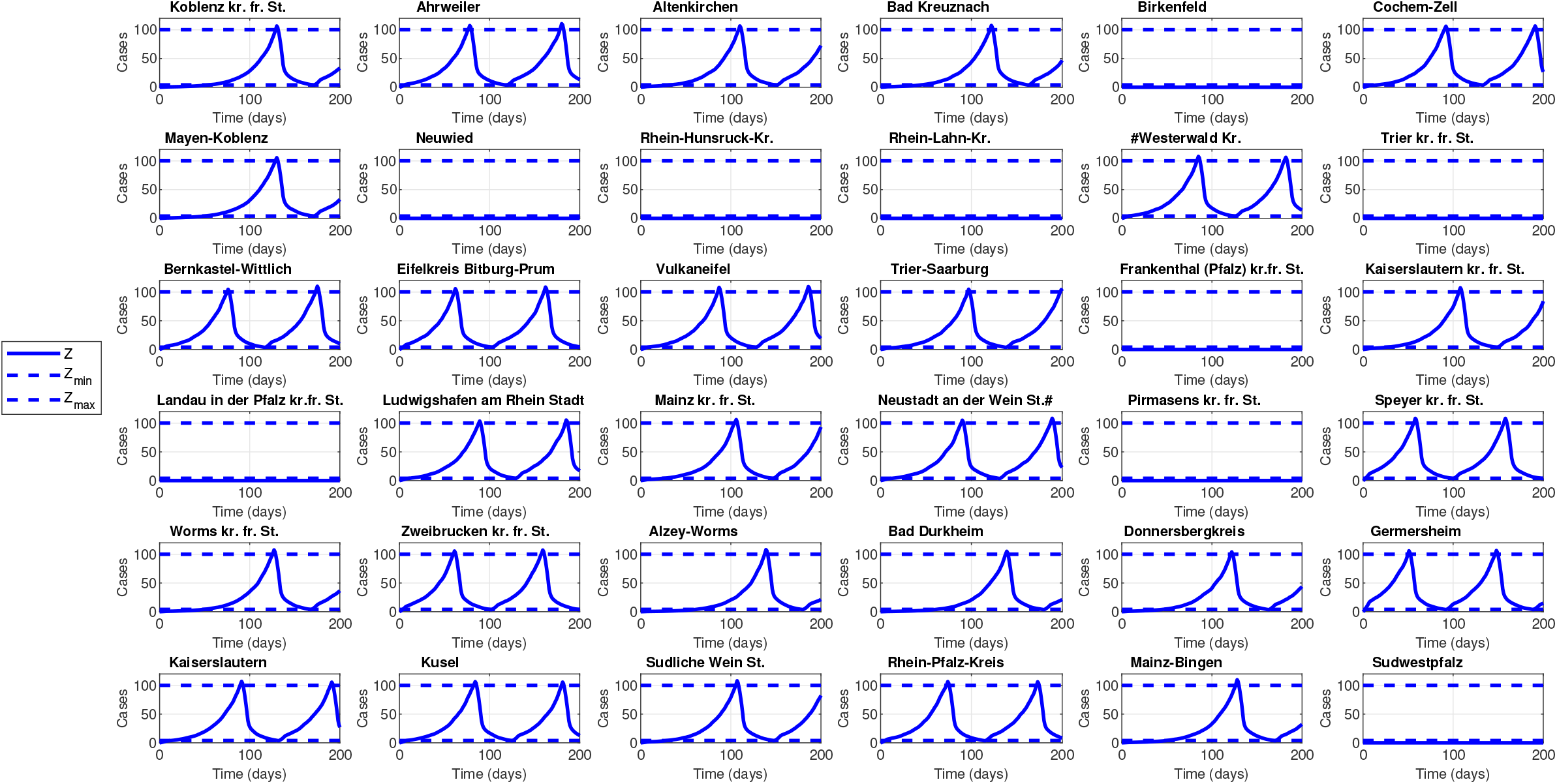
Simulation with no control (*u*_*lock*_ = 0.2) and with no mobility: New cases in the past 7 days per 100,000 inhabitants over time in the 36 counties of RLP with mobility.

**Figure B.14:**
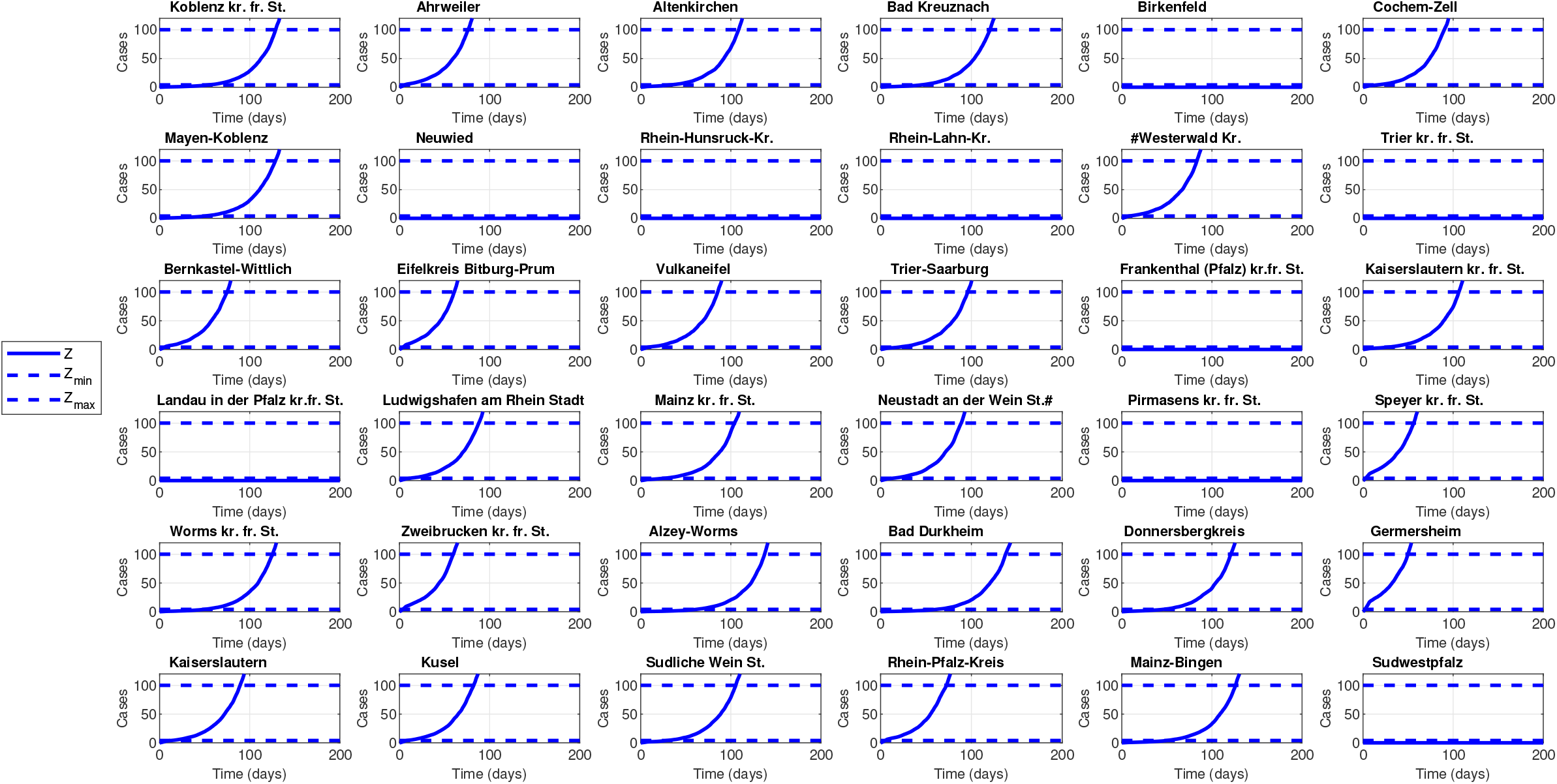
Simulation with no control (*u*_*lock*_ = 1) and with no mobility: New cases in the past 7 days per 100,000 inhabitants over time in the 36 counties of RLP with mobility.

## Notes

### Competing Interest Statement

The authors have declared no competing interest.

### Funding Statement

No funding was received

### Author Declarations

This research was limited to a mathematical data analysis. Therefore, this research would be considered 'minimal risk' and does not come under the definition of research involving human subjects.

